# Neurofeedback-enabled beta power control with a fully implanted DBS system in patients with Parkinson’s disease

**DOI:** 10.1101/2023.05.22.23290293

**Authors:** Manabu Rohr-Fukuma, Lennart H. Stieglitz, Bartosz Bujan, Piotr Jedrysiak, Markus F. Oertel, Lena Salzmann, Christian R. Baumann, Lukas L. Imbach, Roger Gassert, Oliver Bichsel

**Affiliations:** Department of Neurosurgery, University Hospital Zurich, University of Zurich, Switzerland; Clinical Neuroscience Centre, University Hospital Zurich, University of Zurich, Switzerland; Neurorehabilitation, Klinik Lengg, Switzerland; Rehabilitation Engineering Laboratory, Department of Health Sciences and Technology, ETH Zurich, Switzerland; Department of Neurology, University Hospital Zurich, University of Zurich, Switzerland; Swiss Epilepsy Center, Klinik Lengg, Switzerland

## Abstract

Parkinsonian motor symptoms are linked to pathologically increased beta oscillations in the basal ganglia. Studies with externalised deep brain stimulation electrodes showed that Parkinson patients were able to rapidly gain control over these pathological basal ganglia signals through neurofeedback. Studies with fully implanted deep brain stimulation systems duplicating these promising results are required to grant transferability to daily application. In this study, seven patients with idiopathic Parkinson’s disease and one with familial Parkinson’s disease were included. In a postoperative setting, beta oscillations from the subthalamic nucleus were recorded with a fully implanted deep brain stimulation system and converted to a real-time visual feedback signal. Participants were instructed to perform bidirectional neurofeedback tasks with the aim to modulate these oscillations. While receiving regular medication and deep brain stimulation, participants were able to significantly improve their neurofeedback ability and achieved a significant decrease of subthalamic beta power (median reduction of 31% in the final neurofeedback block). We could demonstrate that a fully implanted deep brain stimulation system can provide visual neurofeedback enabling patients with Parkinson’s disease to rapidly control pathological subthalamic beta oscillations.

## 1 Introduction

Recently, neurofeedback (NF) has emerged as a novel approach in the management of neurological and psychiatric disorders (1, 2). It relies on the real-time extraction of relevant features from neuronal activity, that are presented to the subject in real time, who can then develop endogenous techniques to self-regulate this ongoing brain activity. In fact, NF has already been explored for Parkinson’s disease (PD) by numerous studies using electrophysiological and functional imaging methods (3). Most of the electrophysiological NF studies used oscillations of electrical potential in beta frequency (13-35 Hz) as a feedback signal. These oscillations are pronounced in the local field potential (LFP) of the subthalamic nucleus (STN) and internal globus pallidus (GPi) of PD patients (4–7), even though they are not exclusively localised in the basal ganglia, as coupled oscillatory signals are also found in the cortex (6–8).

Beta oscillations can be modulated by PD treatment: Levodopa (4, 9–11) and STN deep brain stimulation (DBS) (12–16) induce a decrease of beta frequency activity in STN and GPi. Hereby, the decline of beta power in the STN induced by levodopa correlates with a decrease in the bradykinesia-rigidity subscore of the Unified Parkinson’s Disease Rating Scale (UPDRS) of the contralateral side (17–19). A correlation of STN DBS induced beta power decrease and motor improvement has also been demonstrated (14, 20). Conversely, there is also evidence that external synchronisation of STN LFP with stimulation at 20 Hz disturbs previously intact motor ability in PD patients (21).

The connection between beta oscillations in the STN and movement is extensively characterised as well. Without dopaminergic medication, voluntary movements lead to a reduction of LFP beta frequency oscillations (5). Particularly, in a study investigating finger tapping movements, intrinsically triggered movements showed a continuous decline of beta power during movement whereas movements triggered through extrinsic signals showed a short-term decrease dependent on the tap onset (22). Interestingly, the decline of beta frequency activity starts prior to the movement (5, 10, 23). This premovement decline is magnified and starts sooner through levodopa administration and correlates with motor ability (10, 23). Due to these results, it was suggested that this decline in beta activity is a correlate of movement preparation (10, 23). Supporting this theory, kinesthetic motor imagery without movement causes a decrease in beta power comparable to performed movements (24).

Having multiple relationships to PD, beta oscillations are suggested as a possible biomarker (25). Indeed, beta oscillations were successfully used as a feedback signal for adaptive DBS in PD patients, thereby achieving greater motor improvement while also relieving battery strain compared to continuous DBS (26). Electroencephalography (EEG) NF training targeting beta bursts, i.e. short-term increases of beta oscillation amplitudes, in the sensorimotor cortex showed a significant decline of beta bursts and faster motor response in healthy subjects (27). Furthermore, there are NF studies in PD patients providing signals through invasive methods. A successful modulation of beta oscillation with a completely implanted electrocorticography (ECoG) electrode over the sensorimotor areas has been demonstrated (28). Most of the NF studies applying invasive methods, though, make use of the electrodes implanted for DBS to provide information about the current state of beta oscillations in the STN. One study showed a significant suppression of beta frequency oscillations in the STN intraoperatively (29). Moreover, three studies were conducted in a postoperative setting making use of the time window between the two operations of DBS electrode implantation and neurostimulator implantation, when the DBS leads were externalised and therefore available for recording (30–32). One study showed a partly significant control over the appearance of beta bursts in the STN (30). Another study of the same group confirmed the significant capability of NF to diminish beta bursts in the STN leading to an improvement in a motor task (31). Moreover, we previously showed that NF using externalised DBS electrodes enabled patients with PD to rapidly gain beta-regulatory control, which in turn quickened movements in a hand pro- and supination task (32).

The experimental setup using externalised leads was valuable to demonstrate the efficacy of DBS NF, but is not suitable for every-day use as the leads ultimately must be internalised. Here, to our knowledge for the first time, we report on NF training with an entirely internalised DBS system to replicate the results of DBS NF studies using externalised DBS electrodes and provide evidence that NF is feasible in a setting similar to daily life.

## 2 Material and methods

### 2.1 Setting and participants

The participants consisted of seven PD patients and one patient with familial PD (five male and three female patients, mean age +/- standard error of mean 68.5 +/- 2.38 years) receiving DBS electrode implantation into the STN. They were numbered with their patient identification (PID) number. A sample size of 6 patients was calculated *a priori* for finding a significant difference between a non-regulating and NF-regulating setting at significance level of α = 0.05/2 and statistical power of 0.8 (33). DBS electrodes were implanted into the STN at the Department of Neurosurgery of the University Hospital Zurich. The recruitment for this study took place postoperatively. The NF trials were conducted on three separate days at the University Hospital Zurich or Klinik Lengg between day 2 and 35 after electrode implantation. Here we report on the outcome of the first day. Medication or stimulation load were not affected by study participation and determined according to the treating neurologists. The post-operative rehabilitation procedure did not differ from that of non-participating patients. The details of the participants are summarised in Table 1.

**Table 1.** Demographic and clinical data of participants. ON and OFF International Parkinson and Movement Disorder Society (MDS) - UPDRS III scores were the result of a preoperative test (34). Levodopa equivalent dose (LED) was calculated using conversion factors from (35, 36). Benserazid and Carbidopa were not considered for calculation (37). Lacking information on the LED of Madopar DR® (Roche Pharma (Schweiz) AG, Basel, Switzerland), its LED was assumed to be equivalent to the one of Madopar HBS® (F. Hoffmann-La Roche, Basel, Switzerland) multiplied with the increased bioavailability of approximately 40% (38). Madopar HBS® was deemed being of controlled release character (39) based on (40). The LED values were rounded two digits before the decimal point.

| PID | age (y) | gender | PD type | side dominance | first manifestation |
| --- | --- | --- | --- | --- | --- |
| 0 | 76 - 80 | female | Morbus Parkinson, akinetic-rigid type | left | 2011 |
| 1 | 56 - 60 | female | familial Parkinson's disease, equivalence type | left | 2013 |
| 2 | 66 - 70 | male | Morbus Parkinson, equivalence type | right | 2013 |
| 3 | 56 - 60 | female | Morbus Parkinson | left | 2008 |
| 4 | 66 - 70 | male | Morbus Parkinson, tremor-dominant type | right | 2015 |
| 5 | 71 - 75 | male | Morbus Parkinson, equivalence type | right | 2011 |
| 6 | 71 - 75 | male | Morbus Parkinson, tremor-dominant type | left | 2004 |
| 7 | 76 - 80 | male | Morbus Parkinson, tremor-dominant type | left | 2014 |

**Table 1. Demographic and clinical data of participants.**
| PID | MDS-UPDRS III<br>OFF/ON | preoperative<br>LED (mg/d) | LED on first session<br>day (mg/d) | stimulation<br>(mA) | postoperative day<br>at first session |
| --- | --- | --- | --- | --- | --- |
| 0 | 46/24 | 1000 | 700 | 0.3 | 9 |
| 1 | 50/23 | 1600 | 1100 | 0.3 | 7 |
| 2 | 29/17 | 1700 | 700 | 1.3 | 30 |
| 3 | 20/10 | 1000 | 600 | 0.3 | 5 |
| 4 | 29/19 | 300 | 0 | 0 | 2 |
| 5 | 78/43 | 2200 | 1600 | 1 | 19 |
| 6 | 50/26 | 3900 | 600 | 1.1 | 17 |
| 7 | 65/29 | 900 | 600 | 1 | 25 |

### 2.2 Surgery and implanted materials

DBS electrode implantation corresponded to the surgical procedure as previously described (32). Leads of the model B33005 SenSight™ (Medtronic, Minneapolis, Minnesota, USA) or 3389 (Medtronic, Minneapolis, Minnesota, USA) were implanted into the STN. The leads were connected via the extensions of the model B34000 SenSight™ (Medtronic, Minneapolis, Minnesota, USA) or 37086 (Medtronic, Minneapolis, Minnesota, USA) to the neurostimulator Percept™ PC B35200 (Medtronic, Minneapolis, Minnesota, USA) during the same surgical procedure or in a second operation on a later date. Information about the DBS system is summarised in Table 2.

**Table 2.** DBS system details and measurement setting.

| PID | implanted lead | utilised extension | utilised neurostimulator |
| --- | --- | --- | --- |
| 0 | Medtronic B3300533 SenSight™ | Medtronic Extension B3400095 SenSight™ | Medtronic Percept™ PC B35200 |
| 1 | Medtronic Lead model 3389-28 | Medtronic Extension model 37086-95 | Medtronic Percept™ PC B35200 |
| 2 | Medtronic B3300533 SenSight™ | Medtronic Extension B3400095 SenSight™ | Medtronic Percept™ PC B35200 |
| 3 | Medtronic Lead model 3389-28 | Medtronic Extension B3400095 SenSight™ | Medtronic Percept™ PC B35200 |
| 4 | Medtronic B3300533 SenSight™ | Medtronic Extension B3400095 SenSight™ | Medtronic Percept™ PC B35200 |
| 5 | Medtronic B3300533 SenSight™ | Medtronic Extension B3400095 SenSight™ | Medtronic Percept™ PC B35200 |
| 6 | Medtronic B3300533 SenSight™ | Medtronic Extension B3400095 SenSight™ | Medtronic Percept™ PC B35200 |
| 7 | Medtronic B3300533 SenSight™ | Medtronic Extension B3400095 SenSight™ | Medtronic Percept™ PC B35200 |

**Table 2. DBS system details and measurement setting.**
| PID | measured STN | electrodes for measurement | beta peak (Hz) |
| --- | --- | --- | --- |
| 0 | right | 8 to 10 | 24.41 |
| 1 | right | 9 to 11 | 27.34 |
| 2 | left | 1 to 3 | 24.41 |
| 3 | left | 1 to 3 | 25.39 |
| 4 | left | 1 to 3 | 24.41 |
| 5 | left | 1 to 3 | 24.41 |
| 6 | right | 9 to 11 | 12.7 |
| 7 | right | 9 to 11 | 8.78 |

### 2.3 Neurofeedback trial

#### 2.3.1 Feedback setting

The participants’ task during the NF trials was to gain control over pathological LFP beta oscillations while receiving visual feedback of their own current beta power in the STN. The fully implanted DBS system provided this information wirelessly over the communicator (model 8880T2, Medtronic, Minneapolis, Minnesota, USA) to the tablet clinician programmer (CT900D, Medtronic, Minneapolis, Minnesota, USA) with the clinician programmer software application (A610 3.0, Medtronic, Minneapolis, Minnesota, USA) visualising the data through BrainSense™ Technology (Medtronic, Minneapolis, Minnesota, USA).

The NF tasks were executed only on the symptom dominant side, therefore the information about the current beta power was only obtained in the STN contralateral to the symptom dominant side. In one case (PID 3), the recording occurred on the STN ipsilateral to the symptom dominant side, as the contralateral STN was not suitable for NF training due to excessive artifacts.

In order to provide visual feedback, a frequency range of locally elevated beta oscillatory activity, referred to as beta peak, was determined in BrainSense™ Survey and selected manually in the BrainSense™ Setup function as the frequency range of beta peaks shows interindividual variability (19, 41). The selected STN side and peak frequency for each patient is listed in Table 2. For PID 6 and 7, peak frequencies in the alpha frequency (8-12 Hz) were selected as no local maximum within the beta frequency was visually identified. It has been hypothesised that when no low beta peak (in the frequency range of 13-20 Hz) is available, it can appear in the high alpha frequency (41). According to the BrainSense™ Technology Summary document, a range of 5 Hz around the manually determined beta peak was used for power calculation. The power of the beta frequency range selected in such way will herein be referred to as beta power.

The bipolar electrode configuration for LFP recording was selected in the Brain-Sense™ Setup function. The two electrodes adjacent to the stimulation electrode (suggested by the program as ‘active therapy’) were chosen for recording (see Table 2). As most of the patients were receiving DBS according to their neurorehabilitation plan, simultaneous stimulation and recording was performed.

Visual feedback was provided by the BrainSense™ Streaming function of the Brain-Sense™ Technology on the tablet Medtronic clinician programmer. Thereby, Brain-Sense™ Technology determined the beta power at a rate of 2 Hz for the selected beta frequency range based on the LFP data (recorded at 250 Hz) and displayed it in a continuous graphical form with time on the x-axis and beta power on the y-axis, as explained in the BrainSense™ Technology Summary document. The default setting of the BrainSense™ Streaming function was not changed. This function displayed two graphs of beta power, one showing the activity of the last approximately 6 s, the other showing the entire streaming session with the last approximately 6 s highlighted in a brighter blue colour. Patients were advised to focus on the brightly highlighted part of the graph displaying the entire streaming session, as this was deemed to provide the best visual information for NF.

#### 2.3.2 Tasks

The main tasks were constructed identical to our previous study (32). The NF trials consisted of three different tasks: baseline, downregulation and upregulation. In the baseline task, baseline activity in the selected beta frequency range was recorded. Participants did not get any visual feedback, were requested to avoid intentionally regulating beta power and refrain from movements. For down- and upregulation tasks, patients received visual feedback. In downregulation tasks, patients were instructed to pay attention to their current beta power and try to lower the curve, i.e. the beta power, by imagining a certain situation, whereas in upregulation tasks, the goal was to drive up the curve. Movements during these tasks were to be avoided by the patients as well. Participants were given an initial strategy to achieve downregulation by imagining asymptomatic movements or situations, and upregulation by imagining situations in which they were afflicted by PD symptoms.

The upregulation part served as the control task for the clinically important downregulation part. This bidirectional NF design has been suggested as a possible control method, as it can expose erroneously achieved NF (42) and was also utilised in two previous DBS NF studies (29, 32).

Furthermore, two blocks without NF were integrated. These blocks each consisted of the three tasks baseline, downregulation and upregulation as well, but the patients did not receive visual feedback for any of these tasks and were asked to regulate the beta power solely by the application of mental strategies. One non-NF block was located before the NF tasks (pre-NF) to record the initial regulatory ability before NF training and one non-NF block after the NF tasks (transfer) to demonstrate how the regulatory ability is modified by the NF training.

#### 2.3.3 Experimental procedure

Each session consisted of five blocks: one pre-NF block, three NF blocks, and one transfer block. Both NF and non-NF blocks were made up of two runs, each consisting of the tasks baseline, downregulation, and upregulation, with each task lasting 60 s. The order of the tasks in the first run of the block was baseline, downregulation, upregulation, whereas the second run had the sequence baseline, upregulation, down-regulation. One BrainSense™ Streaming recording was congruent with one run and therefore had a length of 180 s. All the tasks were performed in sitting position. The experimental procedure is summarised in Table 3.

**Table 3.**
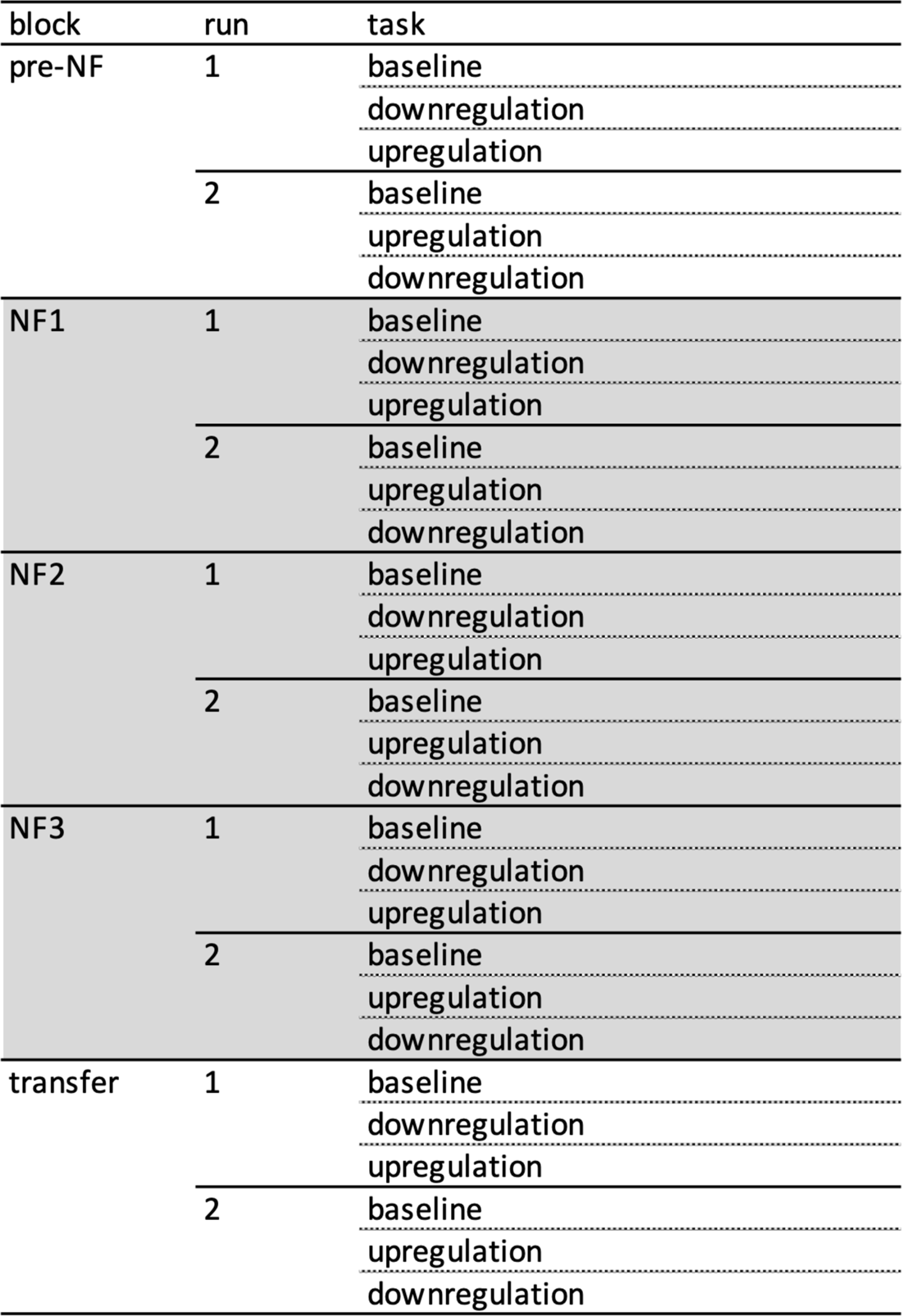
Experimental procedure. A session was defined as the entirety of measurements conducted for one patient on a single day. A session consisted of 5 blocks from which the first and the last block were non-NF blocks. Blocks with NF are highlighted in grey. Each block consisted of 2 runs, of which each contained the tasks baseline, downregulation and upregulation. The sequences of tasks were different in the first and second run of a block.

The described order was chosen to diminish disruptive factors. Changing feedback conditions throughout a run represented one concern. The graph used for feedback displayed the entire beta power of the run and was varying strongly at the beginning of the measurement. To calibrate and provide a stable graph for visual feedback, the baseline task was always put in first place in a run. Moreover, by alternating the order of the down- and upregulation tasks in second and third place, the same visual feed-back condition could be ensured for these tasks in total. The alternating order of down- and upregulation was also important to balance out a carry-over effect (31) as well as the concentration level.

For the first three runs of the session, i.e. the pre-NF block and the first run of the first NF block, mental strategies used for NF were prescribed. For downregulation, participants had to imagine smooth fist forming/opening of the hand contralateral to the recorded STN. The upregulation strategy consisted of a specific self-experienced symptomatic situation. To visualise the neurophysiological mechanisms, a movement-induced beta decline was shown to the patients after the second run. In the subsequent runs, the mental strategy was not predefined, but the patients were encouraged to develop a strategy imagining situations without symptoms for downregulation and situations with symptoms for upregulation.

After the runs in the NF blocks, patients were asked to rate their impression of control they had for the down- and upregulation tasks on a scale from 0 (no ability to control beta power) to 10 (perfect ability to control beta power), herein referred to as agency.

### 2.4 Data and statistical analysis

BrainSense™ Technology stored the session data in a JSON file. The JSON file was then imported to MATLAB (R2021a Update 4, The MathWorks, Inc., Natick, Massachusetts, USA) for analysis, statistical evaluation and graphical illustration. Analysis concerning the difference of beta power between tasks/blocks and the course of beta power during a run was conducted with data stored under the struct ‘BrainSenseLfp’. For the visualisation of the course of beta power during runs (Figure 4), the ‘BrainSenseLfp’ data was normalised by the median beta power value of the baseline task of the same run. The median normalised value of all patients was then determined for every time point of a run, further analysed and graphically illustrated.

To calculate the beta power of the task (Figures 2, 3, 7, 8), the median beta power of each task was determined, and these values were then averaged for the two same tasks of one block (e.g. 2 downregulation tasks in NF1 block). Subsequently, these downregulation values were normalised by the equivalently processed baseline values (Figures 2, 7, 8A) and respective upregulation values by said downregulation values (Figures 3, 8B) of the same block. Statistical analysis in Figures 2 and 3 was conducted with such normalised data. We conducted a paired-sample t-test for evaluating the difference between tasks while the difference between blocks was determined by Wilcoxon signed rank test. Furthermore, linear mixed effects analysis of the relationship between the beta power and the neurofeedback time (pre-NF, NF1, NF2, NF3) was carried out. In order to assess the effect of neurofeedback downregulation time on the beta power, we tested the full model with the effect in question as well as random effect for intercept with subjects as grouping variable against a rudimentary fixed intercept model only. We used a theoretical likelihood test to assess the significance of the random effect.

**Figure 1.**
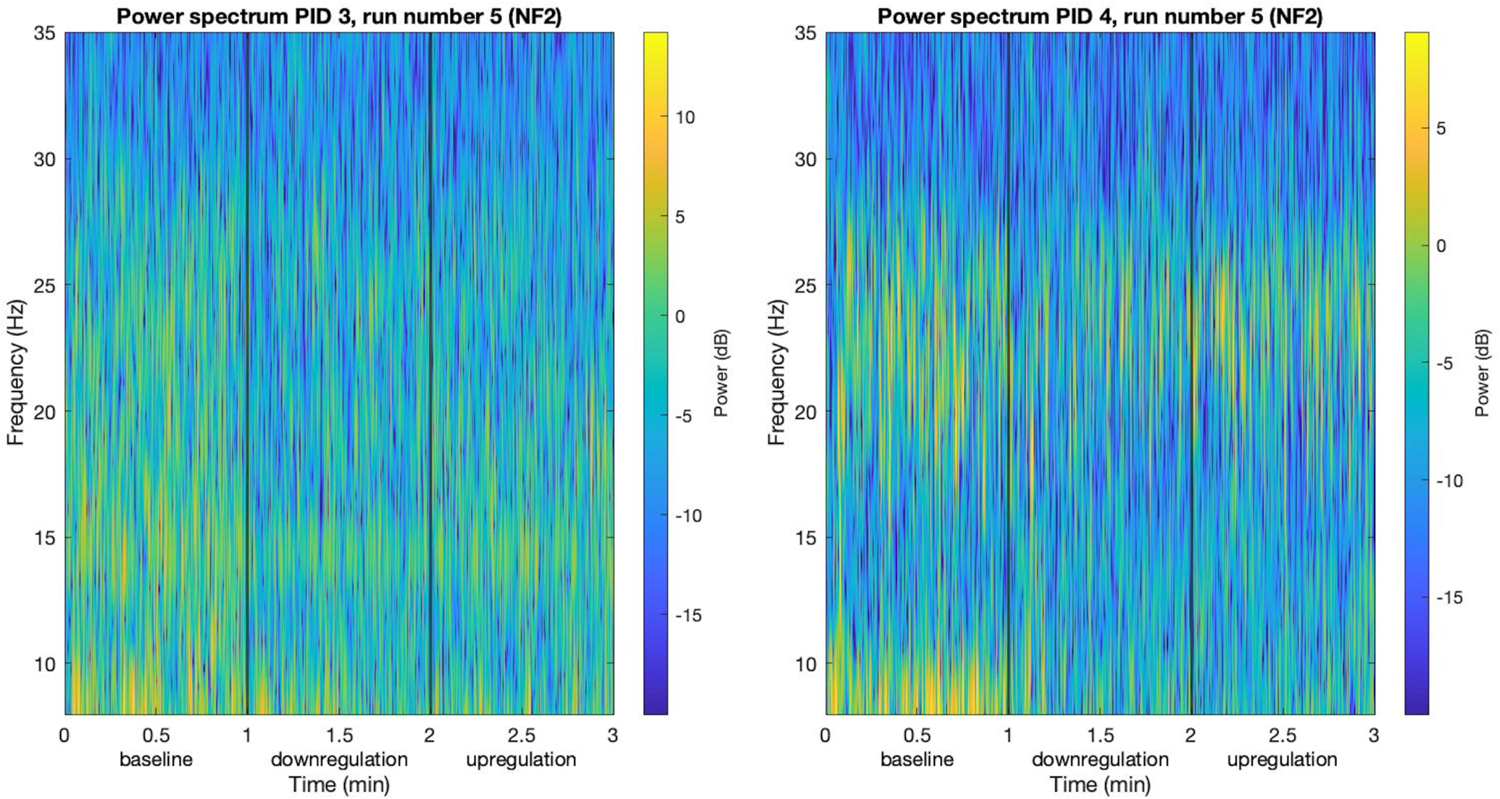
Exemplary runs visualised as power spectra.

**Figure 2.**
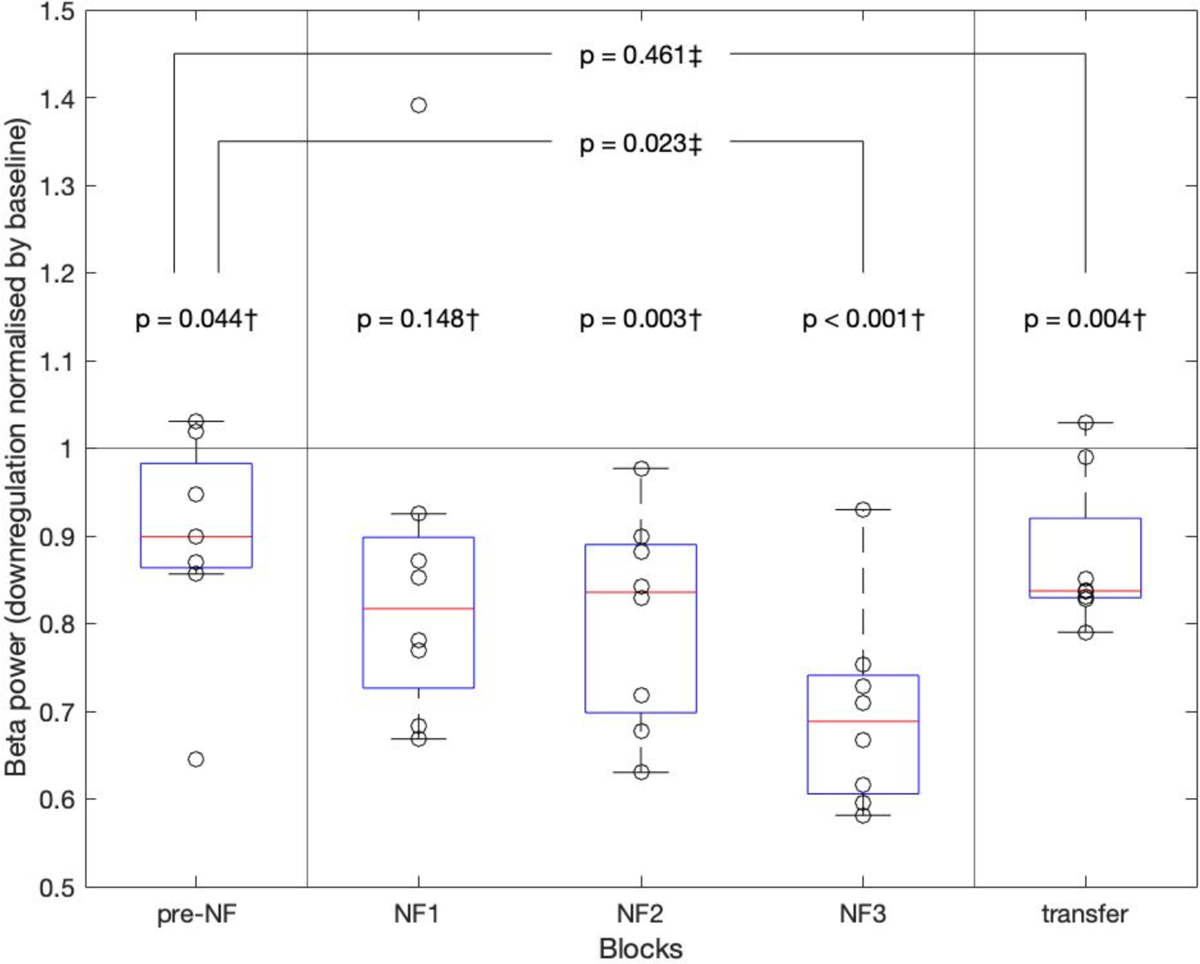
Beta power of downregulation tasks compared with baseline tasks. Median beta power value of downregulation normalised by the median baseline beta power value of the same block visualised as a box- and scatterplot. The black circles of the scatter plot each show the normalised beta power value of each patient. The red line of the boxplot is the median downregulation beta power value of all patients in one block, the blue box displays the interquartile range and the whiskers extend from the nearest edge of the interquartile range box to the maximum or minimum value within the range of 1.5x interquartile range. The line at y=1 corresponds to the baseline. The p-values marked with † were calculated with the paired-sample t-test, comparing normalised downregulation and baseline tasks from the same block. The p-values marked with ‡ were calculated with Wilcoxon signed rank test comparing normalised downregulation tasks from 2 different blocks, connected with lines in the figure.

**Figure 3.**
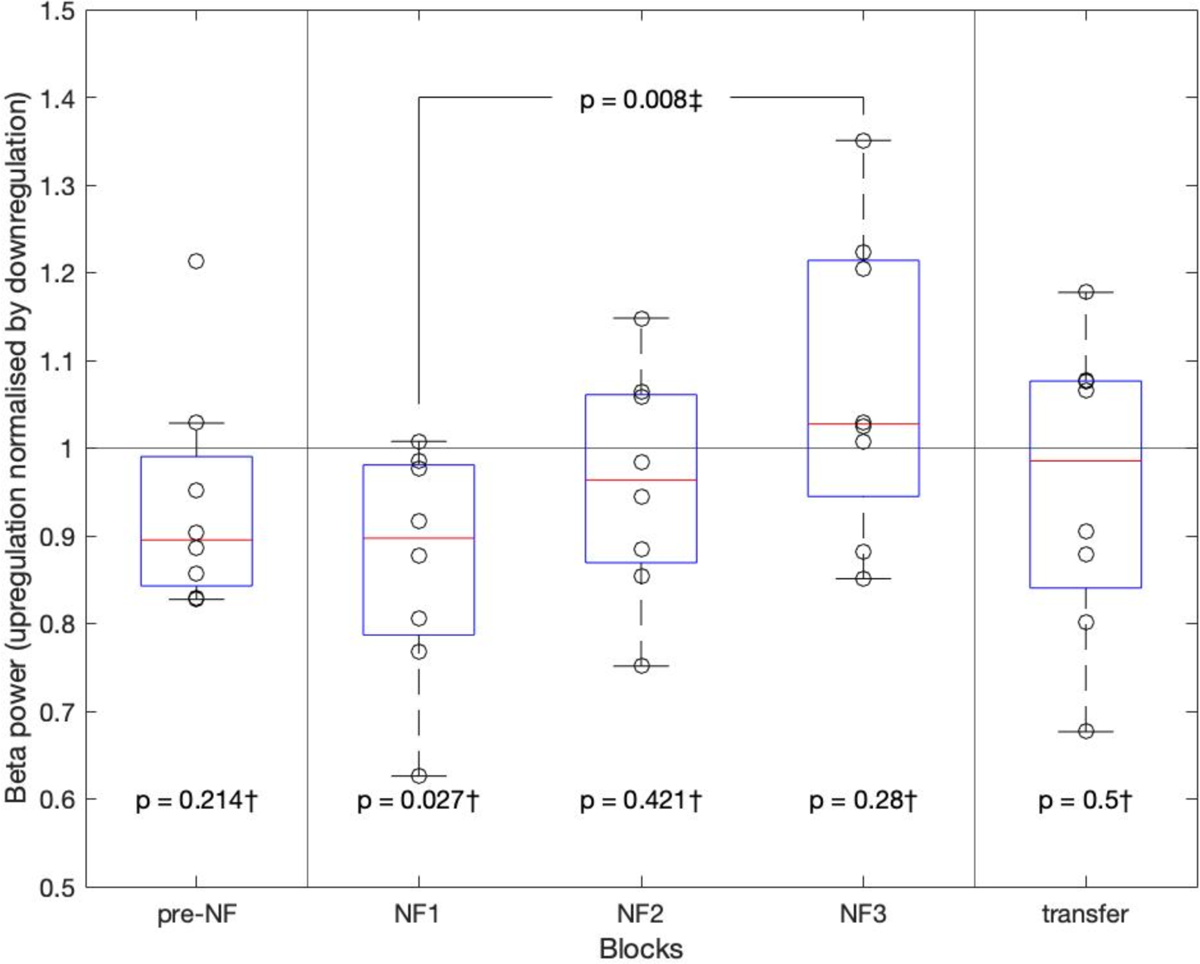
Beta power of upregulation tasks compared with downregulation tasks. Median beta power value of the upregulation block normalised by the median down-regulation beta power value of the same block visualised as a box- and scatterplot. The black circles of the scatter plot each show the normalised value of one patient. The red line of the boxplot is the median value of all patients in one block, the blue box shows the interquartile range, and the whiskers extend from the nearest edge of the interquartile range box to the maximum or minimum value within the range of 1.5x interquartile range. The line at y=1 corresponds to the baseline. The p-values marked with † were calculated with the paired-sample t-test, comparing normalised up- and downregulation tasks from the same block. The p-value marked with ‡ was calculated with Wilcoxon signed rank test comparing normalised upregulation tasks from 2 different blocks, connected with a line in the figure.

**Figure 4.**
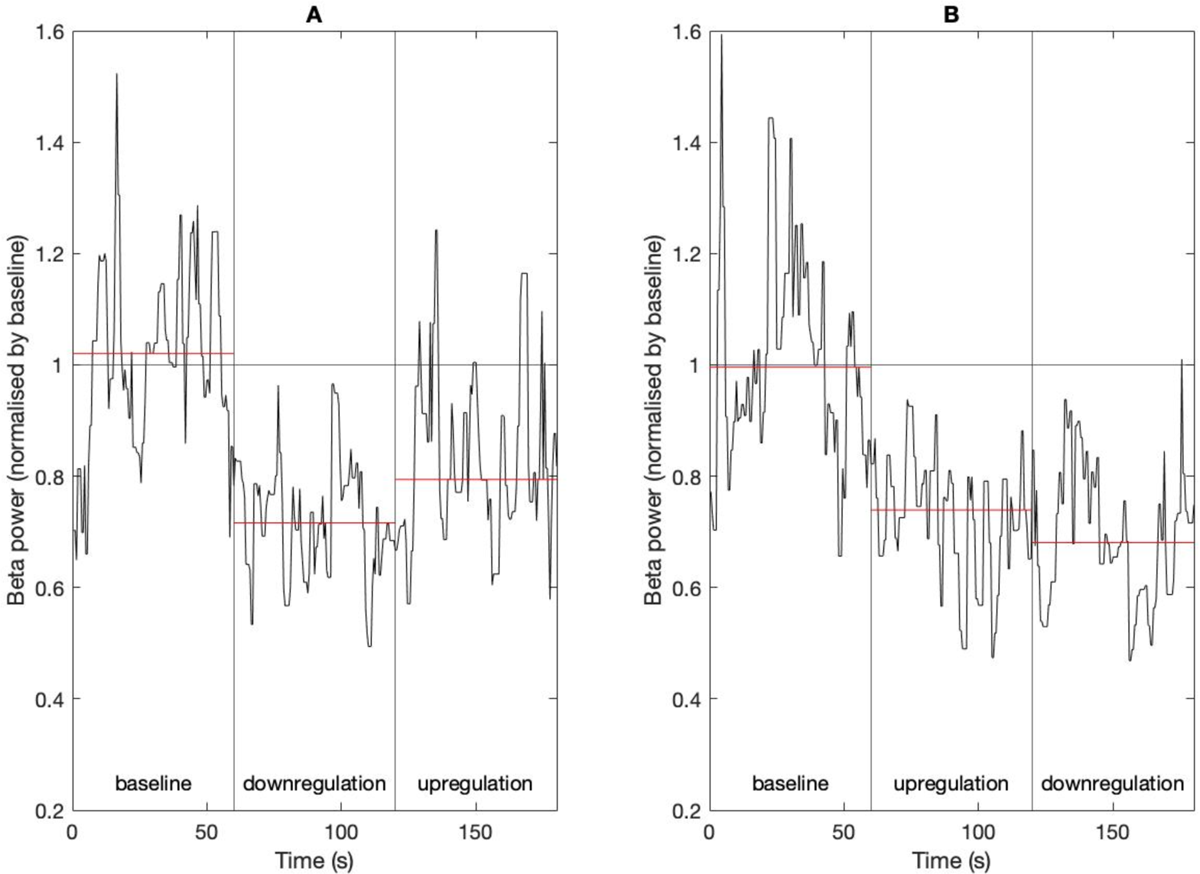
Median beta power in the time course of runs. The median beta power values of NF3 runs of all patients normalised by the median value of the associated baseline tasks are displayed for each time point in NF3 runs as a black graph. The red line shows the median value of the displayed graph in each task. A) shows runs in the order of baseline, downregulation, upregulation, B) shows runs in the order of baseline, upregulation, downregulation.

As two patients were lacking a beta peak after visual inspection and therefore the NF tasks were conducted with a peak located in the alpha band, separate plot visualisations and statistical analyses were carried out excluding these patients (Supplementary Figure 1, 2).

In Figures 7A and 7B, the normalised downregulation values of pre-NF, NF1, NF2, NF3 and transfer were averaged for each person. Influence of number of days since electrode implantation (Figure 7) and agency (Figure 8) were investigated for linear relationship. Hereby, agency values of each task were averaged for one block.

The data struct ‘BrainSenseTimeDomain’ containing the LFP data measured at 250 Hz was used when conducting spectral analysis of power (Figures 1, 5, 6). The basic spectral analysis algorithm of MATLAB, containing fast Fourier transformation, was applied in all baseline tasks for peak analysis (Figure 5), and in baseline and downregulation of all NF tasks for comparison of the power spectrum of such two tasks (Figure 6). The resulting spectral data were averaged for baseline and for downregulation for each patient. Smoothing of the mean spectral data was achieved by calculating the moving average of 250 data points. Local maxima were determined within the frequency range of 13-35 Hz.

**Figure 5.**
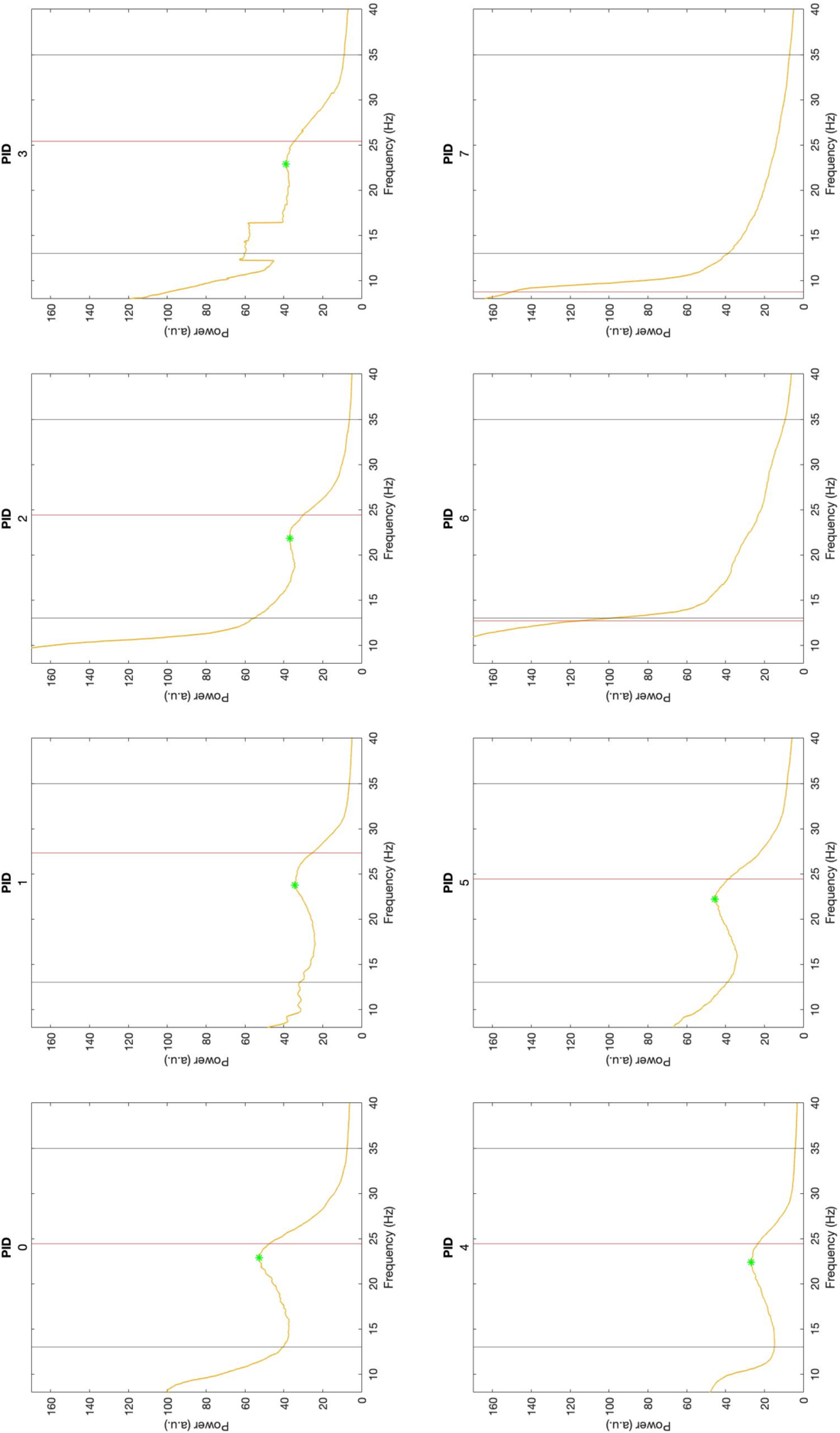
Spectral analysis of all baseline tasks. The yellow line represents the average of the power spectrum of all baseline tasks. The red line represents the frequency, which was manually chosen as peak frequency for feedback in BrainSense™ Setup and the green star shows the beta peak, which manifested as a local maximum in the retrospective analysis. The black lines enclose the beta frequency band defined as 13-35 Hz.

**Figure 6.**
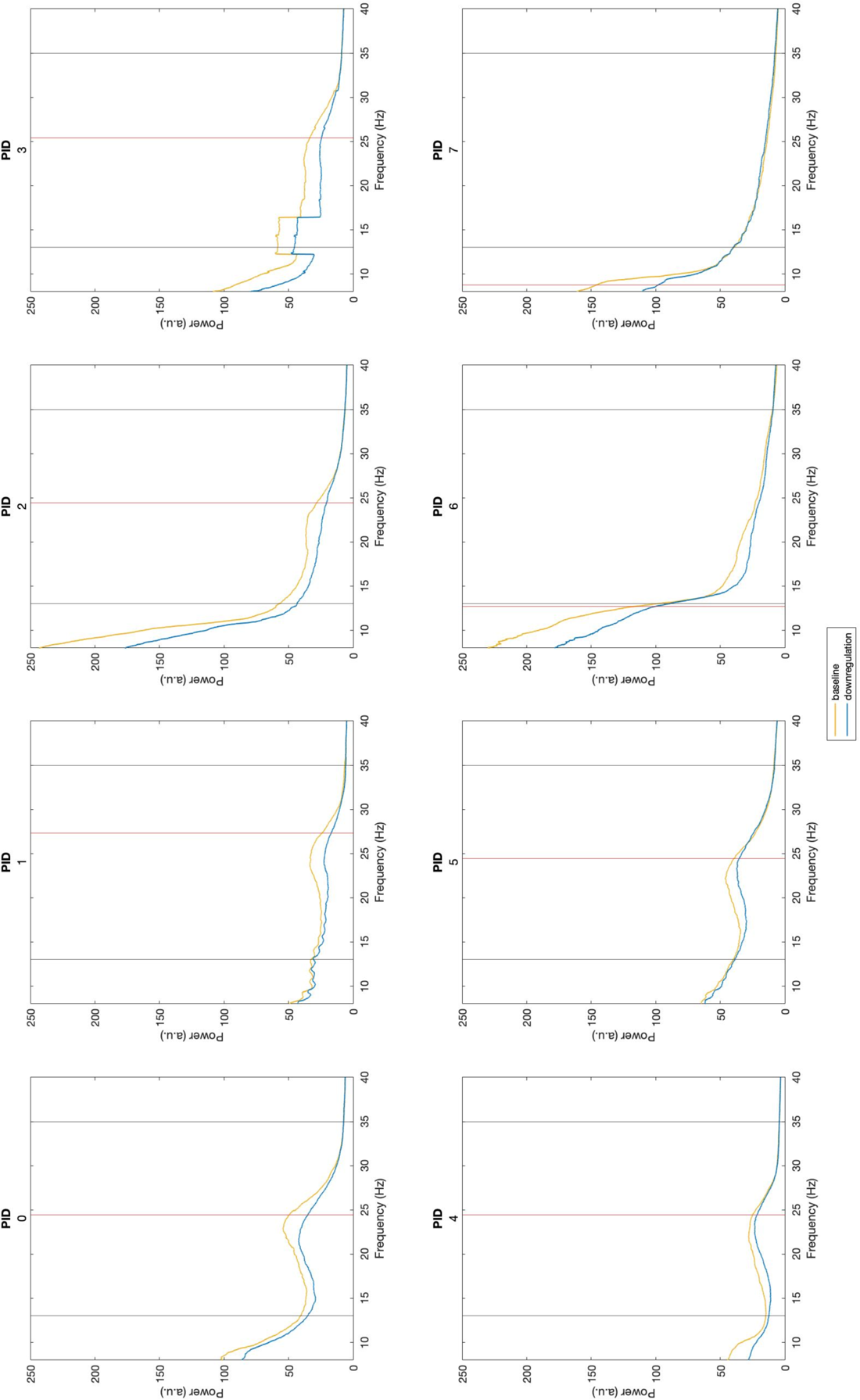
Spectral analysis of NF baseline and downregulation tasks. The yellow line shows the average of the power spectrum of baseline tasks during NF blocks. The blue line displays the average of the power spectrum of downregulation tasks during NF blocks. The red line represents the frequency, which was manually chosen for visual feedback. The black lines enclose the beta frequency band defined as 13-35 Hz.

**Figure 7.**
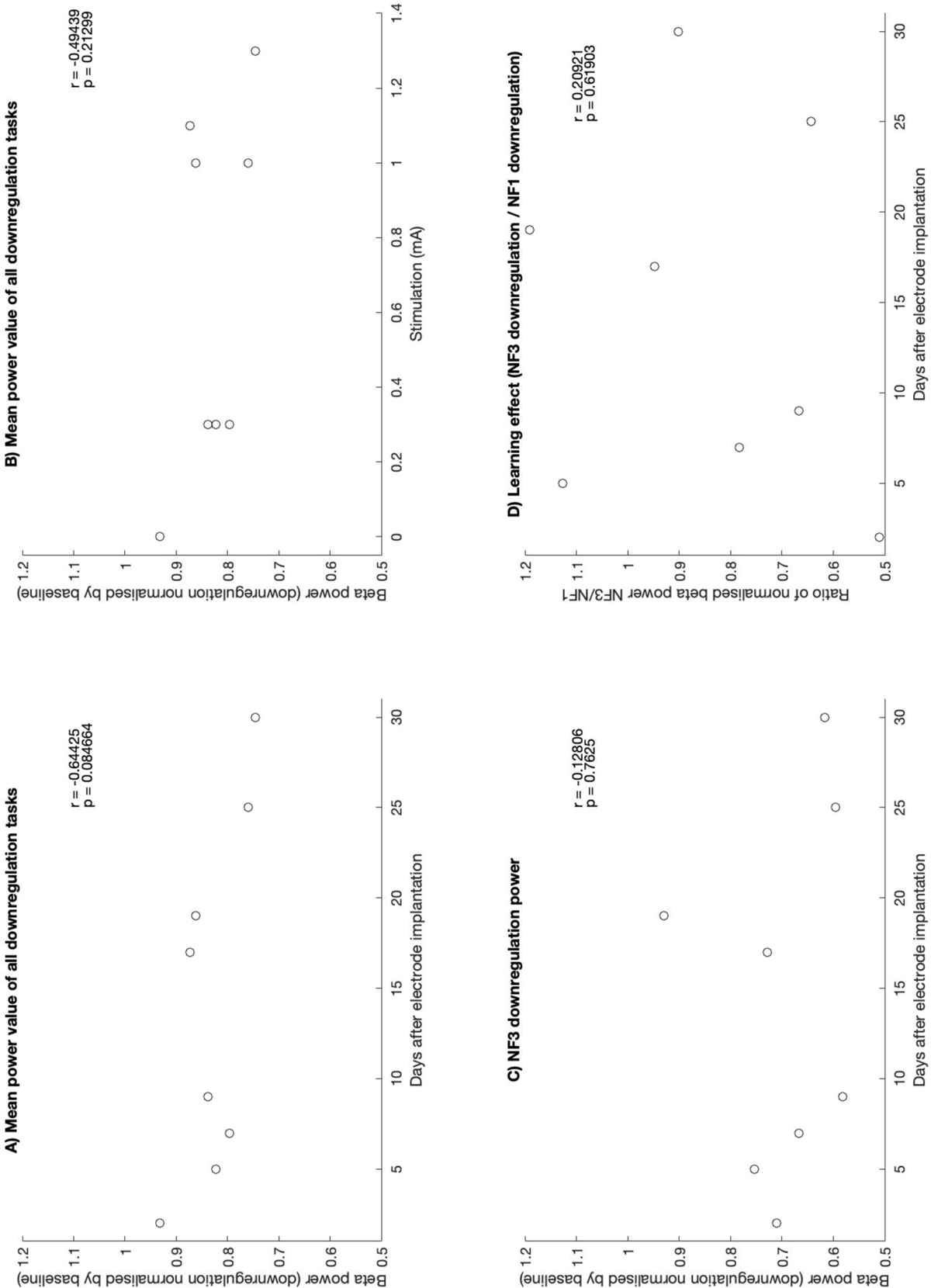
Downregulation depending on time since implantation and stimulation amplitude. The downregulation data was normalised by median baseline values. A & B) The black circles show the average of the normalised median values of all down-regulation tasks for each patient in function of the number of days after electrode implantation (A) or the stimulation amplitude during respective NF sessions (B). C) The black circles show the normalised median values of NF3 downregulation tasks of each patient in function of the number of days after electrode implantation. D) The black circles depict the normalised median values of NF3 downregulation tasks divided by the respective values of the NF1 downregulation tasks for each patient to show the learning effect in downregulation. The r- and p-values of linear correlation were calculated in all schematic representations.

**Figure 8.**
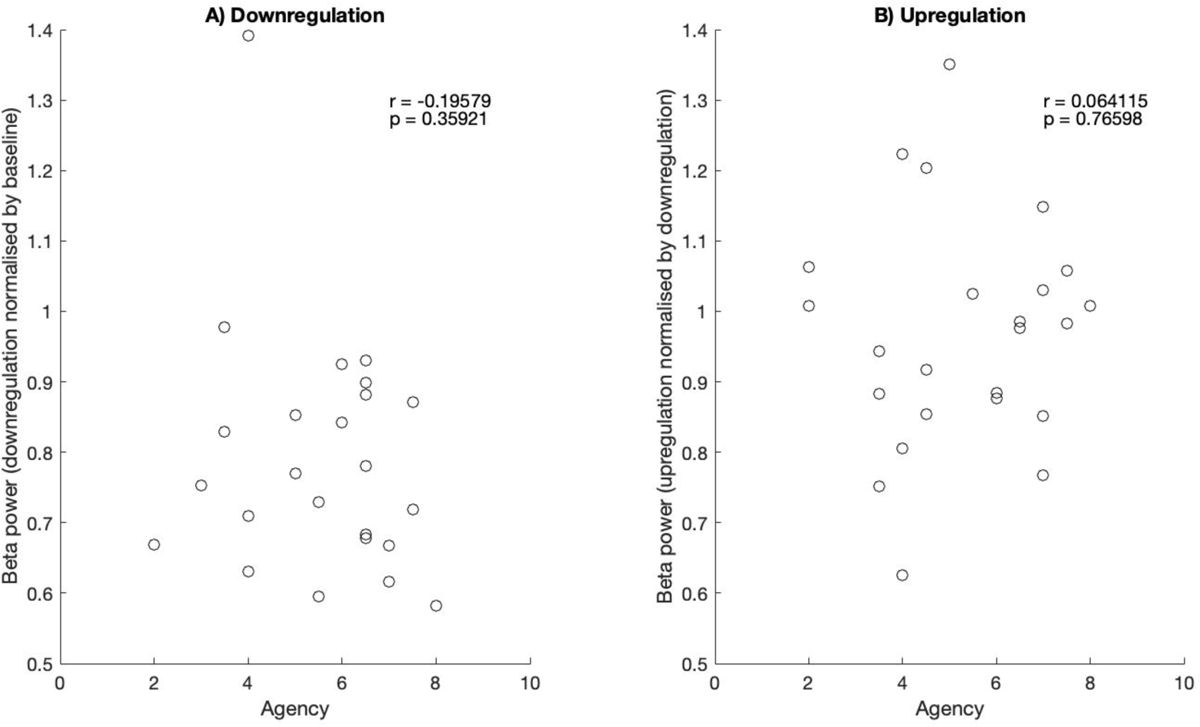
Regulatory capacity in function of agency. Median value of NF downregulation tasks normalised by baseline tasks (A) and NF upregulation tasks normalised by downregulation tasks (B) in dependence on the agency stated by the patients. The r- and p-values of linear correlation are shown.

### 2.5 Ethics

The local ethics committee (Kantonale Ethikkommision Zürich) permitted this study under the BASEC number 2021-00352 and the study was registered under the clinicaltrials.gov number NCT05101161. The study was conducted following the latest version of the World Medical Association Declaration of Helsinki, International Council for Harmonisation of Technical Requirements for Pharmaceuticals for Human Use (ICH) – Good Clinical Practice (GCP) guidelines and local regulations. For the recruitment to the study, an informed written consent was obtained from the patients. This study was conducted in compliance with applicable data protection regulations. Data protection was ensured by encrypting personalised data at acquisition. The unencrypting key as well as unencrypted data were always held at the University Hospital Zurich. Only involved researchers, subjected to professional secrecy, had access to the unencrypted data when and to the extent necessary for the study conduct. In other cases, data evaluation was conducted with encrypted data. The encrypted data can be made available upon reasonable request. Funding for the conduction of this study was provided by the Dr. Wilhelm Hurka Stiftung. There is no conflict of interest to be declared for this study.

## 3 Results

As an exemplary course of measurement, the power spectrum across frequencies against time is graphically illustrated in Figure 1.

Two exemplary runs of two different patients were depicted in a power spectrum over time. The power spectrum for the frequencies between 8 and 35 Hz was visualised for the entirety of the runs of 3 min. Different tasks within a run are separated with a thin black line.

### 3.1 Effect of downregulation compared to baseline

The median beta power values of the downregulation tasks were compared to baseline data of the same block after normalisation. As shown in Figure 2, the median values of the downregulation tasks were below 1 (i.e. below baseline beta power) in all blocks (median of normalised beta power values for pre-NF, NF1, NF2, NF3, transfer: 0.8991, 0.8170, 0.8358, 0.6885, 0.8372). In the NF3 task, a median beta power decrease of 31.15% in downregulations tasks was quantified. There was a significant difference of downregulation to baseline tasks in all blocks (paired-sample t-test) except for NF1, where an outlier influences the result (p-value of pre-NF, NF1, NF2, NF3, transfer: 0.0439, 0.1475, 0.0026, 0.00013, 0.0043). From pre-NF to NF3, the median beta power value of downregulation tasks decreased significantly within minutes of learning (Wilcoxon signed rank test p-value: 0.0234). The median downregulation beta power was lower in transfer compared to pre-NF, however not marking a significant difference (Wilcoxon signed rank test p-value: 0.4609). In the linear mixed effects analysis, the fixed effect of neurofeedback time was significant (p-value: 0.0069) and the introduction of the random effect for intercept with subject as grouping variable significantly improved the model (p-value: 0.0238). Similar results were achieved by excluding the 2 patients without a beta peak (median of normalised beta power values for pre-NF, NF1, NF2, NF3, transfer: 0.9230, 0.8170, 0.7743, 0.6885, 0.8372; paired-sample t-test p-value of pre-NF, NF1, NF2, NF3, transfer: 0.0938, 0.3020, 0.0043, 0.0023, 0.0096), thus showing a significant improvement when comparing the normalised downregulation tasks of pre-NF and transfer blocks (Wilcoxon signed rank test p-value: 0.0313; Supplementary Figure 1).

### 3.2 Effect of upregulation compared to downregulation

Figure 3 shows the median beta power value of the upregulation tasks normalised by the downregulation tasks of the same block. Upregulation had a lower median beta power in all tasks but NF3 (median normalised value of pre-NF, NF1, NF2, NF3, transfer: 0.8953, 0.8972, 0.9636, 1.0276, 0.9855; paired-sample t-test p-value of pre-NF, NF1, NF2, NF3, transfer: 0.2143, 0.0265, 0.4207, 0.2801, 0.4997). For NF1 to NF3, there was a significant increase of upregulation beta power compared to downregulation beta power within minutes of learning (Wilcoxon signed rank test p-value: 0.0078). Excluding the patients lacking a beta peak, similar values were obtained (median normalised value of pre-NF, NF1, NF2, NF3, transfer: 0.8717, 0.8913, 1.0208, 1.0186, 0.9900; paired-sample t-test p-value of pre-NF, NF1, NF2, NF3, transfer: 0.2818, 0.0776, 0.8483, 0.6260, 0.6577; Wilcoxon signed rank test p-value comparing NF1 and NF3: 0.0313; Supplementary Figure 2).

### 3.3 Course of beta power in a run

The median beta power was calculated across all patients for every time point in the NF3 runs and is depicted in Figure 4. It is visible that, in the first few seconds of all tasks the beta power from the earlier task tended to spill over. In NF3, 1 to 7.5 s elapsed until the median beta power (black graph in Figure 4) crossed the median of the median beta power values (red line in Figure 4), calculated for each task, for the first time.

### 3.4 Spectral analysis

All baseline tasks were spectrally analysed to identify the beta peak (Figure 5). There was no clear beta peak within the beta frequency definition of 13-35 Hz for patients with PID 6 and 7. In all other cases, the beta peak defined as a local maximum tended to be lower than the manually chosen peak (mean difference +/- standard error of mean −2.40 +/- 0.28 Hz). The peaks seemed to gather closely in our cohort (mean peak frequency +/- standard error of mean 22.66 +/- 0.28 Hz) The comparison of the spectral analysis of the NF baseline and downregulation tasks showed individual characteristics (Figure 6). It is visible, that there were patients, where only the frequency around the feedback frequency changed (PID 1, 5, 7), one patient, where there was a broad, continuous decrease (PID 3), patients with a decrease at the feedback frequency and a further decrease in another, lower frequency range (PID 0, 2, 4) and even an example of power decrease at a different frequency (PID 6).

### 3.5 Beta power as a function of time since electrode implantation

The dependency of NF regulation ability on number of days since electrode implantation was investigated in our study (Figure 7).

The decrease of power achieved in downregulation compared to the cumulative number of days after electrode implantation showed a tendency of a linear relationship (r = −0.6442, p = 0.0847; Figure 7A).

The same correlation was examined for the relationship of stimulation and beta power decrease through downregulation (Figure 7B). NF sessions took place in the postoperative rehabilitation phase in which the stimulation was slowly increased over the course of days. The magnitude of the stimulation current did not influence the down-regulation ability in the postoperative phase, as it did not show a significant linear relationship (r = −0.4944 p = 0.2130).

The NF3 downregulation ability (Figure 7C) did not hold a significant linear relationship with the number of days after electrode implantation (NF3: r = −0.1281, p = 0.7625).

The learning effect (Figure 7D) defined as the quotient of beta power during NF3 down-regulation tasks and NF1 downregulation tasks revealed no significant linear relationship with the number of days after electrode implantation (NF3/NF1: r = 0.2092, p = 0.6190).

### 3.6 Correlation of patients’ agency and regulation ability

The communicated agency was compared with the effective ability to regulate the beta power (Figure 8). There was neither a significant linear correlation for downregulation (r = −0.1958, p = 0.3592) nor for upregulation (r = 0.0641, p = 0.7660).

## 4 Discussion

### 4.1 Key findings

As the main finding of this study, we were able to show, to our knowledge for the first time, that NF using a *fully implanted* DBS system led to a significant reduction of pathological beta oscillations compared to a non-regulation condition. A significant improvement of downregulation ability within minutes of learning was observed. Although the difference in beta power between upregulation and downregulation tasks remained non-significant, a significant improvement of bidirectional NF control from NF1 to NF3, and, therewith, a significant effect of NF on the regulation of beta oscillations within minutes of learning was demonstrated. This study confirms the results from postoperative NF studies with externalised DBS leads (30–32).

### 4.2 Rapid neurofeedback control over pathological deep brain oscillations using a fully implanted system

As summarised in the Introduction, NF is a possible treatment of PD in addition to the conventional therapies, resulting in several studies in this field. So far, invasive NF was conducted via an implanted ECoG system providing information of the sensorimotor cortex (28) as well as via partially implanted DBS leads (29–32). Herein, we showed that NF regulation is possible with an entirely implanted DBS system and therefore demonstrated its suitability for daily application. This is important as there may be differences in signal quality/resolution and sampling frequency as well as real-time signal processing.

In this study, we provided evidence for a highly significant beta power decrease during the NF downregulation tasks, which is in line with the findings of the other STN DBS NF studies using externalised leads (29–32). It was possible to recreate our previous results (32) in a similar experimental protocol but using a fully implanted DBS system.

### 4.3 The effect of NF on pathological deep brain oscillations in comparison to current treatments

The extent of beta power decrease is also of considerable importance. In the NF3 block, the participants showed a median decrease of 31.15% of beta power in the downregulation tasks compared to the baseline tasks, the greatest value being 41.85%. For comparison, other studies described a beta power decrease of 58.9% (17) and 54.3% (18) with levodopa and 45.8% with DBS (16) without any additional therapy. The result of NF regulation in this study is thus remarkable, as it was achieved under pharmacological therapy and DBS application (note that DBS was most frequently not in the optimal stimulation range, as the study took place during the rehabilitation phase, where DBS was increased slowly over days).

### 4.4 On bidirectional neurofeedback control

Even though the difference of upregulation and downregulation tasks improved significantly from NF1 to NF3, the difference stayed non-significant in absolute terms. The increased challenge of upregulation compared to downregulation was also seen in two other studies implementing upregulation tasks (29, 32). A possible explanation could arise from the use of motor regulation strategies. The decrease of beta activity prior to a movement has been hypothesised to be in the context of movement preparation (5, 10, 23). Furthermore, there is a beta power reduction during kinesthetic motor imagery (24). These findings indicate that downregulation strategies using imagery of asymptomatic movements work so well, as they suit the property of movement preparation and motor imagery while the upregulation strategy, imagining symptomatic movements or situations, might not trigger an increase of beta power. However, in Fukuma et al. (29), where the majority of the participants used strategies other than motor imagery, similar difficulties in upregulation occurred. Therefore, the application of motor regulation strategies might not be the sole explanation of the more challenging nature of upregulation. A further explanation for the difficulties of upregulation might be the imagination of symptoms. Most of the patients were in an oligosymptomatic state when the NF sessions were conducted, which could result in difficulties in imagining symptoms, as symptoms were currently mostly absent. This difference may contribute to the less successful result of upregulation tasks. Ultimately, it might be more difficult to increase an already pathologically increased signal as compared to reducing it. Nonetheless, our findings showing successful bidirectional control of beta power were important to exclude erroneously achieved NF effects (42).

### 4.5 Concerning the choice of the neurofeedback parameter

In the spectral analysis of all baseline tasks, it became visible that not all patients have a beta peak. It was shown that, when withheld from medication, beta peaks are present in the majority of PD patients, whereas beta peak prevalence in the low beta frequency band (defined as 13-20 Hz) decreases to 50% by application of medication (25, 41).

Interestingly, Darcy et al. (41) stated a 100% prevalence of beta peaks in tremor-dominant PD patients withheld from medication in at least one hemisphere, whereas in our study, with all but one patient receiving medication, the 2 patients missing a beta peak on the investigated hemisphere were tremor-dominant PD patients.

The lack of a beta peak in these two tremor-dominant patients can be discussed from three aspects:

Firstly, the frequency definition of the pathological oscillations is inconsistent among studies as some of them include lower frequencies than others. Darcy et al. discussed that the classification of the frequency bands as traditional EEG bands may not suit the parkinsonian electrophysiology, the latter showing a dopamine induced reduction of power in alpha and low beta frequency (41). Furthermore, peaks in the high alpha frequency decrease more strongly by the administration of medication if there is no local maximum in the low beta frequency, and therefore, it has been hypothesised that the low beta peak might also appear in the high alpha frequency (41). In line with these results, it can be observed in Figure 6, that in 4 out of 6 patients showing a beta peak, the reduction in beta power with NF extended into the alpha frequency. Also, the 2 patients without an identifiable beta peak showed a reduction in power of alpha frequencies. Moreover, participants with NF signals in the alpha frequency achieved similar results as patients with feedback markers in the beta frequency. The absence of improvement of the results by excluding the 2 patients lacking the beta peak is another indication that the strict adherence to the beta band may not be of much benefit. It remains to be explored if the localisation of the feedback parameter in the alpha frequency induces a reduction of symptoms through NF.

Secondly, tremor is generally understood as a PD symptom with a spared pathophysiology (25, 43). Remarkably, the correlation of beta power reduction and clinical improvement was only shown for bradykinesia and rigidity (17–19). Furthermore, electrophysiologically, tremor is accompanied by beta power decrease (31, 44–47). Moreover, an increase of oscillations in the STN LFP within the frequency band of 3-7 Hz, described as tremor frequency, was demonstrated (31, 44, 45, 47, 48).

Based on the aforementioned electrophysiological mismatch and finding tremor worsening with NF targeting beta burst, He et al. (31) queried the effectiveness of NF targeting the LFP beta frequency in terms of tremor improvement. This suggestion cannot be evaluated with this study, as the relationship of clinical improvement with NF was not investigated.

Lastly, for one patient lacking a beta peak (PID 6), power decreases different from the feedback frequency were observed in Figure 6. One power decrease was prominentely pronounced in the frequency range of approximately 14 to 24 Hz. This can be interpreted as having missed a possible beta peak, which unveiled itself during the NF training. However, part of this power decreasing band was included in the feedback frequency as well. Furthermore, in Figures 5 and 6, the beta peaks determined in the spectral analysis tended to have a lower frequency than the manually selected peaks. The largest effect of downregulation tended to be at lower frequencies as well. Therefore, it may be useful to adapt the feedback frequency after a few NF regulation tasks to a frequency that is regulated more effectively or that better correlates with clinical improvement.

### 4.6 Towards DBS-neurofeedback in an every-day setting

Previous NF studies with PD patients were restricted to the time window between the two operations while the DBS leads were externalised (30–32), whereas, in this study, the time point of the NF sessions could be selected freely after full implantation of the DBS system. Thereby, downregulation ability may improve with an increasing number of days since the electrode implantation, as we observed a tendency of linear decline of downregulation task beta power in function of the number of postoperative days, as shown in Figure 7A. The increasing stimulation throughout the days after the electrode implantation did not have a clear effect on the downregulation ability, as there was no significant correlation of stimulation amplitude and downregulation ability, which can be observed in Figure 7B. Nevertheless, the observation from Figure 7A does not mean that NF sessions should be carried out at later stages of the postoperative phase, as the learning effect, calculated as the quotient of NF3 and NF1 downregulation beta power, was not influenced by the number of days since electrode implantation. However, further studies might demonstrate superior results when the tasks are conducted at later stages of the rehabilitation phase.

The agency stated by the patients at the end of a NF run failed to show a correlation with the actual regulation result. As the self-assessment of regulation ability was not accurate, it might be difficult for patients to efficiently pursue effective regulation strategies in an entirely independent NF training. Therefore, it could be beneficial for patients to start NF trials in a supervised setting with instructions and advices.

### 4.7 Limitations and outlook

As previously shown (32), the ability to downregulate beta power via NF and an improvement of this ability within a short time span was demonstrated in this study as well. With the analysis conducted in this study, no conclusion can be drawn for an effect over multiple days. As this study demonstrated the feasibility of NF control with an entirely implanted DBS system, it will be possible to investigate NF with this experimental setup over a much larger time span in the future.

In this study, muscle contractions were not controlled with electromyography (EMG) electrodes. This could represent a possible disruptive factor, as voluntary movements induce a decrease in beta frequency activity (5). However, movements were prohibited as a regulation strategy and the examinators visually controlled for overt movements. We waived an EMG control, as it was shown in previous studies, that patients did not tend to use muscle contraction in NF settings and that the beta activity decrease of NF was not induced erroneously through movement (29–31).

As, within a run, the transition to the next task followed without a pause, the experimental results were affected by a spill-over effect. Analysing the course of runs, the spill-over effect varied between 1 and 7.5 s. A carry-over effect was described in the work of He et al. (31). Excluding the spill-over effect, the beta power regulation could reveal to be even more effective. However, it can still be stated that the NF effect of modulating beta power is deployed relatively quickly, within seconds.

The lack of beta peaks in some patients can be discussed as a limiting factor, as NF can possibly not be provided in all PD patients. However, by selecting a peak in lower frequencies, the modulatory ability of the oscillatory activity in the STN was similar to when the selected peak was in the beta frequency. This can also be seen in the similarity of results when the patients lacking a beta peak were excluded (Supplementary Figure 1, 2). The clinical effect of NF in the alpha frequency band, though, remains to be investigated.

The aim of this study was to provide evidence of NF feasibility with an entirely implanted DBS system and therewith the suitability of NF in an every-day setting. The effect of NF in a situation of symptom exacerbation, which was suggested as a possible utilisation of NF (32), was not evaluated in this study, as most of the patients were oligosymptomatic. Furthermore, motor behavioural outcome was not investigated in this study, whereby other NF studies provide evidence of clinical improvement (31, 32). Finally, by showing the feasibility of NF with an entirely implanted DBS system, longterm studies as well as testing in symptom exacerbating situations become feasible and are of great interest.

## 5 Conclusion

We demonstrated successful and significant regulation of STN beta band power through NF using an entirely implanted DBS system in patients suffering from PD. Patients learned to control pathological deep brain oscillations within a few minutes of NF training. Of note is the high amount of beta power regulation (median decrease of 31.15%, maximum decrease of 41.85% in the last NF block) achieved using NF despite concurrent treatment with medication and DBS. As NF was conducted with an entirely implanted DBS system, the evidence presented in this study encourages the transfer of DBS-NF to the every-day setting and the conduction of long-term NF studies.

## Data Availability

The encrypted data can be made available upon reasonable request.

