## Supplementary Figure 1 and 2 for "Neurofeedback-enabled beta power control with a fully implanted DBS system in patients with Parkinson’s disease"

### Supplemental Data

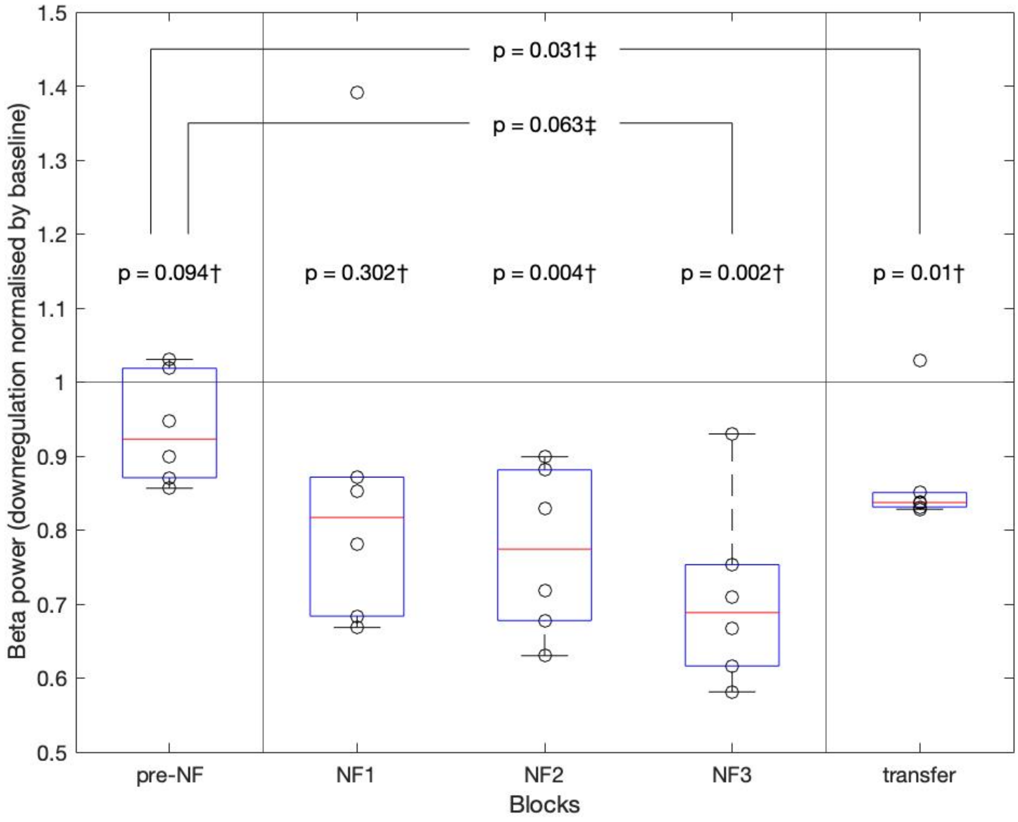


**Supplementary Figure 1. Beta power of downregulation tasks compared with baseline tasks excluding patients without a beta peak.** Median beta power value of downregulation normalised by the median baseline beta power value of the same block visualised as a box- and scatterplot. The 2 patients lacking the beta peak were excluded for this plot and statistical analysis. The black circles of the scatter plot each show the normalised beta power value of one patient. The red line of the boxplot is the median downregulation beta power value of all patients in one block, the blue box displays the interquartile range and the whiskers extend from the nearest edge of the interquartile range box to the maximum or minimum value within the range of 1.5x interquartile range. The line at y=1 corresponds to the baseline. The p-values marked with † were calculated with the paired-sample t-test, comparing normalised downregulation and baseline tasks from the same block. The p-values marked with ‡ were calculated with Wilcoxon signed rank test comparing normalised downregulation tasks from 2 different blocks, connected with lines in the figure.

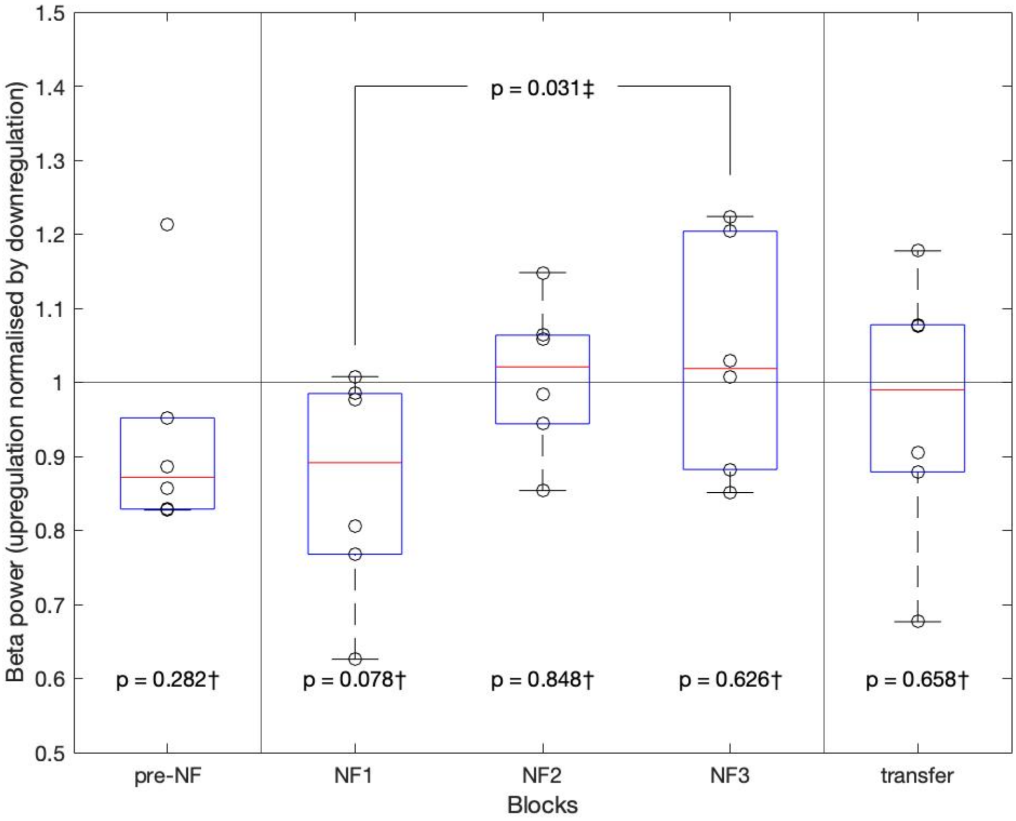


**Supplementary Figure 2. Beta power of upregulation tasks compared with downregulation tasks excluding patients without a beta peak.**

Median beta power value of the upregulation block normalised by the median downregulation beta power value of the same block visualised as a box- and scatterplot. The 2 patients lacking the beta peak were excluded for this plot and statistical analysis. The black circles of the scatter plot each show the normalised value of one patient. The red line of the boxplot is the median value of all patients in one block, the blue box shows the interquartile range, and the whiskers extend from the nearest edge of the interquartile range box to the maximum or minimum value within the range of 1.5x interquartile range. The line at y=1 corresponds to the baseline. The p-values marked with † were calculated with the paired-sample t-test, comparing normalised up- and downregulation tasks from the same block. The p-value marked with ‡ was calculated with Wilcoxon signed rank test comparing normalised upregulation tasks from 2 different blocks, connected with a line in the figure.
